# Risk of Parkinson’s Disease in Stroke Survivors: A Nationwide Cohort Study in South Korea

**DOI:** 10.1101/2023.06.02.23290911

**Authors:** Hea Lim Choi, Jong Hyeon Ahn, Won Hyuk Chang, Wonyoung Jung, Jinyoung Youn, Dong Wook Shin

**Affiliations:** Department of Family Medicine & Supportive Care Center, Samsung Medical Center, Sungkyunkwan University School of Medicine, Seoul, South Korea; Department of Neurology, Samsung Medical Center, Sungkyunkwan University School of Medicine, Seoul, South Korea; Neuroscience Center, Samsung Medical Center, Sungkyunkwan University School of Medicine, Seoul, South Korea; Department of Physical and Rehabilitation Medicine, Center for Prevention and Rehabilitation, Heart Vascular Stroke Institute, Samsung Medical Center, Sungkyunkwan University School of Medicine, Seoul, South Korea; Department of Family Medicine, Kangdong Sacred Heart Hospital, Hallym University, Seoul, South Korea; Department of Clinical Research Design and Evaluation, Samsung Advanced Institute of Health Science and Technology (SAIHST), Sungkyunkwan University, Seoul, South Korea; Department of Digital Health, Samsung Advanced Institute of Health Science and Technology (SAIHST), Sungkyunkwan University, Seoul, South Korea

**Keywords:** Stroke, Parkinson’s disease, disability, nationwide, cohort

## Abstract

**BACKGROUND:** Previous studies have examined the risk of stroke in patients with Parkinson’s disease (PD), but the incidence of PD onset among stroke survivors and its risk according to severity of post-stroke disabilities is scarcely investigated. This study aims to determine whether the risk of PD is increased among stroke survivors using a retrospective cohort with a large population-based database.

**METHOD:** We used data collected by the Korean National Health Insurance Service from 2010–2018 and examined 307,361 stroke survivors and 380,917 sex-and age-matched control subjects to uncover the incidence of PD. Cox proportional hazards regression was used to calculate the hazard ratio (HR) and 95% confidence interval (CI), and the risk of PD was compared according to presence and severity of disability.

**RESULTS:** During 4.31 years of follow-up, stroke survivors had a 1.67 times higher risk of PD compared to matched control subjects (adjusted HR, 1.67; 95% CI, 1.57–1.78). The risk of PD was greater among stroke survivors with disabilities than among those without disabilities, even after adjustment for multiple covariates (adjusted HR, 1.72; 95% CI, 1.55– 1.91 and adjusted HR, 1.66; 95% CI, 1.56–1.77, respectively).

**CONCLUSION:** Our study demonstrated increased risk of PD onset among stroke survivors. Health professionals need to pay careful attention to detecting movement disorders as clues for diagnosing PD.

## INTRODUCTION

Stroke is the second leading cause of death,^1^ and the burden of stroke has increased with its contribution to long-term disability. Stroke and Parkinson’s disease (PD) are common problems in the aged population, and the co-existence of these disorders can lead to particularly unfavorable outcomes.^2, 3^ Previous studies have reported that PD patients with a history of stroke have increased risk of hospitalization,^4, 5^ and even a silent vascular burden can affect the motor and non-motor symptoms of PD patients.^6^ Therefore, the relationship between PD and stroke could be clinically important.

A case–control study^7^ performed in the United Kingdom with 3,637 PD cases and age-and sex-matched controls showed that stroke was more prevalent in PD cases compared to the control group (adjusted odds ratio [OR], 1.65; 95% confidence interval [CI], 2.00–2.47). Similar results were found in smaller studies from the United Kingdom^8^ (crude OR, 2.2; 95% CI, 1.1–4.6) and China (OR, 2.58–6.77).^9^ A retrospective cohort study^10^ enrolling a Korean population also found a greater risk of ischemic stroke in 5,259 PD patients compared to age-and sex-matched controls (adjusted hazard ratio [HR], 3.88; 95% CI, 3.17–4.75).

Development of stroke in PD patients was explained by (1) their similar etiology, which is common among the elderly^11–13^; (2) involvement of increased reactive oxygen species that damage dopaminergic neurons and promote atherosclerotic changes; and (3) shared risk factors such as hypertension, diabetes mellitus, and dyslipidemia.^14–17^

However, most previous studies examining the association between PD and stroke investigated stroke risk in PD patients, and there are limited studies that have directly investigated the incidence of PD among stroke patients (Supplementary Table S1).

Additionally, involuntary movement disorders including parkinsonism and the sequelae in poststroke patients may interfere with the diagnostic process of PD. Therefore, we designed a retrospective cohort study using a large population-based database to investigate the risk of PD development in stroke survivors.

## METHODS

### Data Sources and Study Setting

This retrospective cohort study used data from the Korean National Health Insurance Service (KNHIS) database, a single national insurer covering approximately 97% of the Korean population that collects all medical information on patients, such as visits to medical facilities, prescriptions, and diagnoses recorded as International Classification of Disease, 10^th^ revision (ICD-10) codes. The remaining 3% of the Korean population is covered by a Medicaid program funded by the government, although their records also are managed by the KNHIS.

The KNHIS provides a biennial health screening program to all employees regardless of age or adults who are older than 40 years old.^18^ This program provides extensive information including anthropometric characteristics (blood pressure, height, weight, etc.), health behaviors (regular exercises, smoking status, alcohol consumption, etc.), and laboratory results (blood glucose, lipid profiles, urine analysis). The data obtained from this program have been widely used in various epidemiological studies.^19^

### Study Subjects

This study included 800,646 patients diagnosed with stroke between January 1, 2010, and December 31, 2018. The definition of stroke was based on the recording of ICD-10 codes (I60-I64: includes both ischemic [I63, I64] and hemorrhagic stroke [I60, I61, I62]) with a claims code of brain resonance imagining or brain computed tomography.^20–22^ These stroke patients were randomly matched 1:1 based on age and sex to a control group, with matching performed by year so that stroke survivors diagnosed in a specific year were matched with control subjects alive in that same year. Among the participants, we included those who participated in the national health checkup program within 2 years from the index year and excluded those who were aged <40 years (n=6,719), had a previous history of traumatic brain injury (n=1,039), had a diagnosis of PD prior to stroke diagnosis (n=2,898), or were missing information (n=17,372). This resulted in a total of 307,361 stroke survivors. After applying the same inclusion and exclusion criteria to the matched control group, a total of 380,917 controls was selected for this study. The process of patient selection is illustrated in Figure 1.

**Figure 1.**
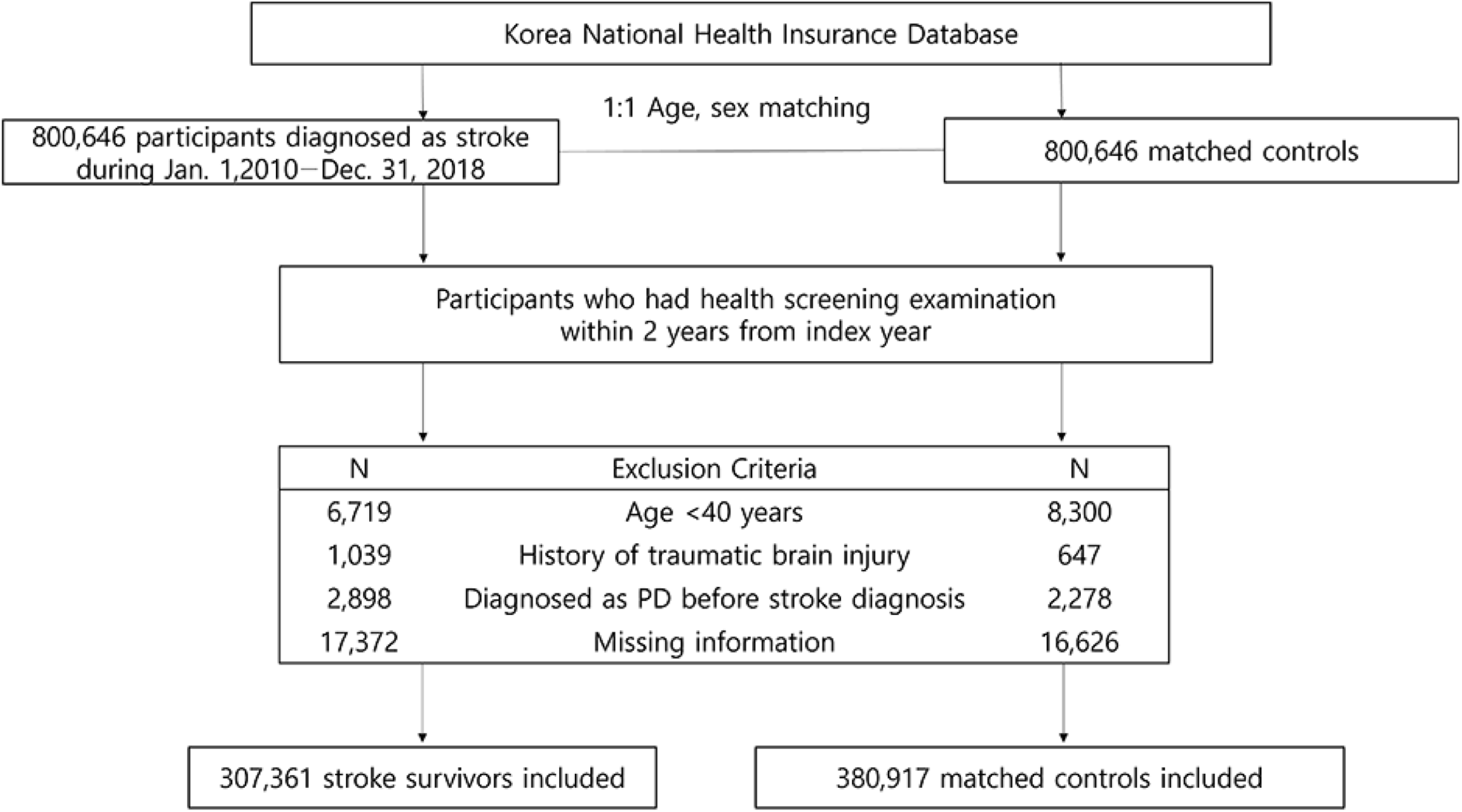
Flowchart of the study enrollment process.

This research was approved by the Institutional Review Board (IRB) of Samsung Medical Center (IRB no. 2020-12-068), and the requirement for informed consent from participants was waived due to the nature of de-identified retrospective data.

## Severity of Disability After Stroke

Severity of disability after stroke was determined according to disability grades defined by the Korea National Disability Registration System (KNDRS).^23, 24^ Fifteen types of disability have been legally defined by the KNDRS, although the degree of disability was graded from 1 (most severe disability) to 6 (mild disability) during the study period. In the present study, disability was graded as severe or mild. As people with disability can obtain social benefits once registered with the KNDRS, almost all eligible people apply for registration. To be registered, people with a disability need to submit certified proof from a specialized physician who determines the severity of their disability based on the criteria provided by the government. In the case of stroke, people who met the criteria can apply to KNDRS for “disability due to brain injury” 6 months after the date of stroke as their disease status became stable. Such patients should be assessed by neurologists, neurosurgeons, or physiatrists to determine of disability severity using the modified Barthel score, and we dichotomized grades 1–3 as severe disability and grades 4–6 as mild disability for analysis^25^ (Supplementary Table S2).

### Study Outcome and Follow-up

The primary outcome of the study was newly diagnosed PD, which was identified by both ICD-19 code (G20) and registration code (V124) of the Rare Intractable Disease management program in Korea, as in prior studies.^26–28^ Patients diagnosed with PD were identified with code V124, which is specified by criteria comparable to the UK Parkinson’s Disease Society Brain Bank Diagnostic Criteria,^29^ and the diagnosis is confirmed by a neurologist or neurosurgeon. The study subjects were followed from the date of first diagnosis of stroke or matched index date for the matched control group to the date of newly diagnosed PD or the end of the follow-up period (December 31, 2019), whichever came first.

### Covariates

We defined low household income as the lowest 20% of insurance premiums, which refers to the proxy for insurance demand, and categorized the place of residence into metropolitan, urban, and rural areas. Smoking status was divided into current, former, and never-smoker groups. Heavy alcohol consumption was defined as >30 g of alcohol per day. Participants who exercised ≥5 times a week at a moderate intensity for ≥30 min or 3 times a week at a high intensity for ≥20 min were defined as regular exercisers. Body mass index was calculated as weight in kilograms divided by height in meters squared. Fasting glucose and total cholesterol levels were measured after 8 h of fasting. Information on comorbidities was obtained from claims data (ICD-10 codes) and prescription information before the index date. Diabetes mellitus was defined as E10–14 and more than one claim for anti-diabetic medication, hypertension was defined as I10–11 and more than one claim for antihypertensive medication, while dyslipidemia was defined as E78 and more than one claim for prescribing lipid-lowering medications. To assess the overall comorbidity load, the Charlson comorbidity index (CCI) was used.^30^

### Statistical Analysis

Descriptive figures were presented as mean ± standard deviation for continuous variables or number (%) for categorical variables. Kaplan–Meir analysis was conducted to show the incidence probabilities of PD in stroke survivors compared to matched controls. We calculated the HR and 95% CI for PD using Cox proportional hazards regression. In the multivariate model (Model 3), we adjusted variables related to PD development, such as age, sex, socioeconomic variables (household income and place of residence), health behaviors (smoking status, alcohol consumption, and regular exercise), comorbidities (type 2 diabetes, hypertension, and dyslipidemia), and CCI score.^31^ Moreover, we compared the risk of PD according to presence and severity of disability. Statistical analysis was conducted using SAS version 9.4 (SAS Institute, Cary, NC, USA), and *P<*0.05 was considered statistically significant.

## RESULTS

### Baseline Characteristics of the Study Population

Table 1 describes the baseline characteristics of the study population. In the stroke survivor group, the mean age was 66.7 years and 56.26% were men. Stroke survivors had lower household incomes, were more often current smokers, and more frequently consumed alcohol compared to matched controls. A lower proportion of stroke survivors performed regular exercise and had higher body mass indices, systolic and diastolic blood pressures, fasting glucose, and total cholesterol before stroke diagnosis compared to matched controls. Higher percentages of comorbidities of type 2 diabetes (31.01%), hypertension (77.91%), and dyslipidemia (68.54%) were found among stroke survivors, and thus the CCI score. The mean follow-up duration was 4.31 ± 2.73 years for stroke survivors and 5.30 ± 2.52 years for matched controls.

**Table 1.** Baseline Characteristics of the Study Population.

| **Variables** | **Matched controls**<br>**(n=380,917)** | **Stroke survivors**<br>**(n=307,361)** | ***P* value** |
| --- | --- | --- | --- |
| Age (years) | 66.5 ± 11.3 | 66.7 ± 11.1 | <0.001 |
| Sex (male) | 217,295 (57.05) | 172,926 (56.26) | <0.001 |
| Income (lowest 20%) | 72,072 (18.92) | 66,782 (21.73) | <0.001 |
| Place of residence (urban) | 163,631 (42.96) | 119,285 (38.81) | <0.001 |
| Current smoker | 62,563 (16.42) | 73,107 (23.79) | <0.001 |
| Heavy alcohol consumption | 24,152 (6.34) | 23,346 (7.6) | <0.001 |
| Regular exercise | 82,808 (21.74) | 55,020 (17.9) | <0.001 |
| Body mass index (kg/m<sup>2</sup>) | 24.06 ± 3.08 | 24.15 ± 3.23 | <0.001 |
| Systolic blood pressure (mmHg) | 127.35 ± 15.34 | 131.88 ± 17.40 | <0.001 |
| Diastolic blood pressure (mmHg) | 77.31 ± 9.81 | 79.98 ± 11.20 | <0.001 |
| Fasting glucose (mg/dL) | 103.78 ± 26.31 | 110.53 ± 38.17 | <0.001 |
| Total cholesterol (mg/dL) | 194.49 ± 38.56 | 197.08 ± 41.3 | <0.001 |
| Comorbidities |  |  |  |
| Type 2 diabetes | 72,613 (19.06) | 95,310 (31.01) | <0.001 |
| Hypertension | 200,742 (52.7) | 239,470 (77.91) | <0.001 |
| Dyslipidemia | 138,929 (36.47) | 210,657 (68.54) | <0.001 |
| Charlson comorbidity index | 1.86 ± 1.92 | 4.54 ± 2.46 | <0.001 |
| Follow-up duration (years) | 5.3 ± 2.52 | 4.31 ± 2.73 | <0.001 |

Data are presented as mean ± standard deviation or number (%).

### Risk of PD in Stroke Survivors Compared to Matched Controls

Incidence probability of PD was higher among stroke survivors than that of matched controls (log-rank *P<*0.001). Adjusted HR (aHR) for PD during the follow-up period among stroke survivors was 1.67 (95% CI, 1.57–1.78) compared to matched controls (Table 2, Figure 2). The risk of PD was higher for both stroke survivors with disabilities (aHR, 1.72; 95% CI, 1.55–1.91) and those without disabilities (aHR, 1.66; 95% CI, 1.56–1.77). Stroke survivors with mild disability showed higher aHR of 1.82 (95% CI 1.62–2.05) than those with severe disability (aHR, 1.50; 95% CI, 1.25–1.79) compared to the matched controls.

**Figure 2.**
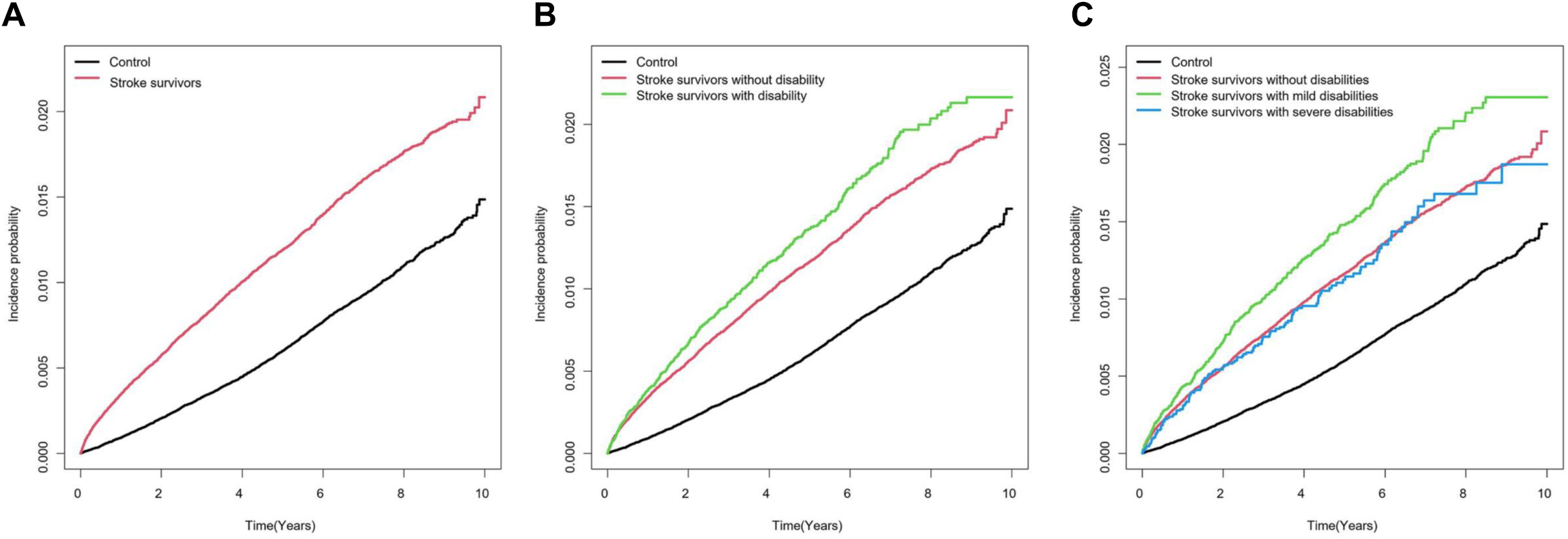
Kaplan-Meier curves of estimated incidence probability for risk of Parkinson’s disease. (**A**) Risk of Parkinson’s disease in stroke survivors and control group. (**B**) Risk of Parkinson’s disease in stroke by presence of disability. (**C**) Risk of Parkinson’s disease in stroke by severity of disability.

**Table 2.** Hazard Ratios and 95% Confidence Intervals for Incidence of Parkinson’s Disease Among Stroke Survivors Compared to the Matched Control Group.

|  | Subjects<br>(n) | Events<br>(n) | Follow-up<br>duration<br>(person-years) | IR | Model 1<br>HR (95% CI) | Model 2<br>HR (95% CI) | Model 3<br>HR (95% CI) |
| --- | --- | --- | --- | --- | --- | --- | --- |
| **Comparison with the matched control group** |  |  |  |  |  |  |  |
| Matched controls | 380,917 | 2,600 | 2,018,389.5 | 1.3 | 1 (Ref.) | 1 (Ref.) | 1 (Ref.) |
| Stroke survivors | 307,361 | 3,174 | 1,323,560.8 | 2.4 | 1.86 (1.76–1.96) | 1.57 (1.48–1.67) | 1.67 (1.57–1.78) |
| **Comparison according to presence of disability** |  |  |  |  |  |  |  |
| Matched controls | 380,917 | 2,600 | 2,018,389.5 | 1.3 | 1 (Ref.) | 1 (Ref.) | 1 (Ref.) |
| Stroke survivors |  |  |  |  |  |  |  |
| No disability | 269,400 | 2,688 | 1,145,846.3 | 2.3 | 1.82 (1.72–1.92) | 1.57 (1.47–1.66) | 1.66 (1.56–1.77) |
| Disability | 37,961 | 486 | 177,714.5 | 2.7 | 2.12 (1.92–2.34) | 1.63 (1.47–1.81) | 1.72 (1.55–1.91) |
| **Comparison according to severity of disability** |  |  |  |  |  |  |  |
| Matched controls | 380,917 | 2,600 | 2,018,389.5 | 1.3 | 1 (Ref.) | 1 (Ref.) | 1 (Ref.) |
| Stroke survivors |  |  |  |  |  |  |  |
| No disability | 269,400 | 2,688 | 1,145,846.3 | 2.3 | 1.82 (1.72–1.92) | 1.56 (1.47–1.66) | 1.66 (1.56–1.77) |
| Mild disability | 25,260 | 353 | 119,738.9 | 2.9 | 2.29 (2.05–2.56) | 1.72 (1.53–1.93) | 1.82 (1.62–2.05) |
| Severe disability | 12,701 | 133 | 57,975.7 | 2.3 | 1.78 (1.49–2.12) | 1.44 (1.21–1.73) | 1.50 (1.25–1.79) |

Abbreviations: IR, incidence rate per 1,000 person-years; HR, hazard ratio; CI, confidence interval.
Model 1: Unadjusted.
Model 2: Adjusted for age, sex, and Charlson comorbidity index.
Model 3: Adjusted for Model 2 + socioeconomic variables (household income and place of residence), smoking status, alcohol consumption, regular exercise status, and comorbidities (type 2 diabetes, hypertension, and dyslipidemia).

## DISCUSSION

To the best of our knowledge, this is the first large population-based study to investigate the incidence of PD among stroke survivors and to demonstrate that PD risk was 1.7 times higher in stroke survivors than matched controls.

The mechanism of developing PD after a stroke event needs to be elucidated but can include cerebral ischemia-induced aggregation of alpha-synuclein (α-syn), and loss of dopaminergic neurons can lead to PD development. Aggregated α-syn and loss of substantia nigra dopaminergic neurons were observed in rodents with infarction.^32^ In parallel with this result, a comparative study of human red blood cells demonstrated increased level of the oligomeric form of α-syn in both ischemic stroke and PD patients compared to normal participants.^33^

Second, shared risk factors in stroke and PD can be attributed to PD onset in stroke patients as well as development of stroke in PD patients. Known stroke risk factors such as age, hypertension, diabetes, heart failure, and obstructive sleep apnea were reported to increase the risk for PD,^34^ suggesting similar pathophysiology between the two disorders.^35^ While we adjusted all available health behavioral and comorbidity factors, there might be a pathophysiological link between stroke and PD that was not fully addressed by our multivariate model.

Additionally, shared genetic factors can explain the increased PD risk in stroke patients. A genome-wide association study^36^ revealed that stroke and PD share five common genes (*PX7, LBH, ZCCHC10, DENND2A*, and *NUDT14),* which were expressed differentially in stroke and PD patients compared to healthy subjects. These studies help to understand the causal relationship between stroke and PD and to elucidate the mechanism to support our results.

Last, structural brain lesions from stroke could have lowered the threshold of PD symptoms by altering the “motor reserve.” Previous models explain the discrepancy between the clinical and pathological severity of PD using the concept of “motor reserve.” Motor reserve in PD may compensate for the dopaminergic degeneration in PD, and parkinsonian symptoms can present when motor reserve fails to compensate for the dopaminergic degeneration in PD patients. Motor reserve in PD is associated with various brain structures, including the basal ganglia and extra-basal ganglia structures, and brain structural lesions from stroke can damage the motor reserve in PD.^37–39^ In our study, PD was more common in stroke survivors; thus, our results supported the connection between PD and stroke.

In terms of disability from stroke, PD was more common in stroke survivors with disability. Considering various brain structures associated with poor motor reserve, more structural lesions or larger brain area involvement could be related to the development of PD. However, when we divided participants into stroke patients with mild or severe disabilities, PD was more common in those with a mild disability compared to those with a severe disability. Even though we also discussed various possible mechanisms that cause PD in stroke patients, parkinsonism could be masked by severe disability from stroke. Because PD is a neurodegenerative disease, development of parkinsonism can be insidious and slowly progressive. If stroke survivors already show severe disability such as hemiparesis or limitations in daily activity, they may face barriers to health care access, and detection of parkinsonism by a physician or caregivers might not be easy. Thus, the diagnosis rate could be lower in stroke survivors with severe disabilities than in those with mild disabilities.

Our study has important clinical implications. It would be detrimental for stroke survivors to develop PD in terms of morbidity and mortality. Previous studies have reported that stroke patients with PD experience longer hospitalization stays; higher rates of morbidities such as pneumonia, sepsis, and acute kidney injury; and increased mortality rates compared to those without PD.^5, 40^ Deteriorated movements due to PD can negatively affect the outcome of rehabilitation in stroke patients.^41^ Therefore, clinicians should be encouraged to screen for symptoms of PD in stroke survivors to achieve early diagnosis, provide timely dopamine replacement therapy, and introduce early rehabilitation to stroke survivors with disabilities to maintain quality of life.

There are several limitations in our study. First, a chance to diagnose PD instead of vascular parkinsonism should be considered—it has been reported that the rate of misdiagnosis of PD accounts for approximately 15%–30% of cases.^42^ Our study minimized this error by excluding patients with secondary parkinsonism (G21), including vascular parkinsonism (G21.4). Last, since emerging data suggest ethnic variations in the incidence and manifestations of PD,^43–45^ our result should not be generalized as we considered only the Korean population.

## CONCLUSION

In conclusion, this nationwide population-based cohort study demonstrated that stroke survivors have an increased risk of developing PD. Future studies are needed to further elucidate the underlying mechanism of PD onset in stroke patients and to observe their association in various ethnic groups.

### Non-standard Abbreviations and Acronyms

KNHIS: Korean National Health Insurance Service
KNDRS: Korea National Disability Registration System
CCI: Charlson comorbidity index
α-syn: alpha-synuclein

## Data Availability

The data that support the findings of this study are available from the corresponding author, [DWS], upon reasonable request.

## Acknowledgments

None

## Supplemental Material

Table S1. Previous Studies of the Association Between Stroke and PD

Table S2. Definitions of Disability Grade after Brain Injury

